# Administration of Normal Saline versus Ringer’s Lactate in Critically Ill Patients: A Protocol and Statistical Analysis Plan for a Secondary Analysis of the FLUID trial (FLUID-ICU)

**DOI:** 10.64898/2026.01.16.26344276

**Authors:** Lauralyn McIntyre, Dean Fergusson, Deborah Cook, Alison Fox-Robichaud, Michael Pugliese, Monica Taljaard

## Abstract

**Introduction:** Recent evidence from randomized controlled trials and meta-analyses in the critically ill suggest small but clinically important differences in death and requirement for renal replacement therapy that favor Ringer’s lactate as compared to normal saline. To futher contribute to this evidence base, we will perform a secondary analysis of the FLUID trial with a focus on critically ill patients (FLUID – ICU). The FLUID trial was hospital wide cluster randomized cross-over trial that compared normal saline to Ringer’s lactate.

**Objectives:** The primary objectives of FLUID-ICU are to examine the effect of Ringer’s lactate compared to normal saline compared to on the primary composite outcome and secondary outcomes in critically ill patients. As secondary exploratory objectives, we will evaluate the primary composite outcome and secondary outcomes within prespecified subgroups defined by age, sex, case mix group, Elixhauser Co-morbidity Index, presence of infection with organ dysfunction, traumatic brain injury, and trauma with an injury severity score of >= 12.

**Methods:** FLUID was an open-label two-period, two-sequence, cross sectional, cluster-randomized cross over trial that was conducted in 7 hospitals in the province of Ontario, Canada. Hospitals were assigned to either Ringer’s lactate or normal saline for a period of 12 weeks and after a washout period, switched to the other fluid at 12 weeks. The primary composite outcome was death or re-admission to hospital within 90 days after the index admission. Secondary outcomes were individual components of the primary outcome as well as length of stay in hospital, an incident emergency department visit post hospital discharge, initiation of dialysis within 90 days after the index admission, and discharge to a facility other than home. All data were obtained from health administrative databases.

In FLUID-ICU, we will examine the primary and secondary outcomes in a cohort of critically ill patients, defined as patients admitted to the ICU within 1 day after admission to hospital. A sensitivity analysis will relax this definition to 2 days after admission to hospital. The analyses will be performed at the individual patient level using random effects logistic regression with adjustment for prespecified covariates.

**Ethics and Dissemination:** The results of FLUID-ICU will be published in a peer-reviewed journal. As a secondary analysis of anonymized data, this study does not require research ethics board approval. All data is obtained from a population based health administrative database (ICES) in the province of Ontario, in aggregate and without patient identifiers.

## Introduction

Ringer’s lactate (RL) and Normal saline (NS) have been administered as resuscitation fluids for several decades. RL and other balanced crystalloid fluids contain less chloride than NS and reduce the risk of hyperchloremic acidosis.^1^ Some observational studies in the critically ill and surgical patient populations also suggest that balanced crystalloids as compared to NS may reduce the risk of death and requirement for renal replacement therapy.^2-5^ Several recent randomized controlled trials (RCTs) comparing balanced crystalloids to NS in critically ill patients have been conducted.^6-10^ A systematic review^11^ of 35,884 participants in 13 critical care trials comparing saline to balanced crystalloids found no significant difference in mortality (28.2% versus 27.9%; relative risk 0.96, (95% CI 0.91-1.01)), or in the use of renal replacement therapy (13.2% and 12.7% respectively; relative risk 0.95 (95% CI, 0.81 to 1.0)^11^. However, this systematic review^11^ and another individual-patient level meta-analysis^12^ using a Bayesian analytic approach concluded that there is a high probability that balanced crystalloids are associated with reduced in-hospital mortality, and a lower use of renal replacement therapy.

Recently, our team conducted the FLUID trial which was a hospital-wide 43,626 patient cluster randomized cross over trial in 7 centres in the province of Ontario, Canada that compared RL to NS on a primary composite outcome of death or re-admission to hospital within 90 days of the index admission.^13^ Clinical data were collected from provincial health administrative data in Ontario. In contrast to RCTs in the critically ill, the FLUID trial enrolled patients hospital-wide, addressing an important evidence gap by evaluating the effect of these two crystalloid fluids from first entry to the hospital until hospital death or discharge. Notably, individual patient exposure to fluids was unknown in this cluster randomized cross-over trial; however, hospital wide adherence (measured using logistical services reports) was 78.2% versus 93.6% in the RL versus NS arms. Although FLUID found no statistically significant differences between RL and NS in the primary composite outcome measured as a hospital-level proportion (adjusted mean difference: -0.53%; 95% Confidence Interval: -1.85 to 0.79), or in any of the secondary outcomes, the accumulation of evidence from recent RCTs in the critically ill suggest that balanced crystalloids as compared to NS may result in small but clinically relevant reductions in serious adverse events (death and requirement for renal replacement therapy)^11-12^.

In this study - FLUID-ICU - we propose a secondary analysis of the primary composite outcome and secondary outcomes in the FLUID trial among critically ill patients. Based on prior evidence from RCTs and meta-analyses, we hypothesize that the direction of the effect for the primary composite outcome will favor RL over NS^6-10,11,12^. As secondary exploratory objectives, we aim to evaluate the primary composite outcome and secondary outcomes within prespecified subgroups defined by age, sex, case mix group, Elixhauser Co-morbidity Index, presence of infection with organ dysfunction, traumatic brain injury, and trauma with an injury severity score of >= 12.

## Methods

### Design

FLUID-ICU is a pre-planned nested cohort study and subgroup analysis of the main FLUID trial. FLUID was an investigator-initiated open-label pragmatic two-period two-sequence cross-sectional cluster randomized cross-over trial conducted at 7 academic and community hospitals in Ontario, Canada between August 2016 and March 2020. There was no individual patient recruitment; all clinical data were obtained from health administrative sources housed at ICES (formerly known as the Institute for Clinical Evaluative Sciences) which is an independent, nonprofit research institute whose legal status under the provincial health information privacy law allows the collection and analysis of health care and demographic data. Clusters were defined as hospital sites, allocated to one of two sequences.

### Eligibility Criteria for Clusters and Participants

Clusters were eligible for FLUID if they admitted at least 1500 participants per study period and had level II (support for failure of one organ, non-invasive ventilation, or post operative recovery) or III (support for multiple failed organs, advanced or prolonged mechanical ventilation support) ICU capacity according to Critical Care Services Ontario. All index admissions over the study period were eligible for inclusion, regardless of exposure to the study fluid (which was not available in the provincial health administrative database at the patient level). An index admission was defined as a patient’s first hospital admission with no prior hospital admission in the previous 90 days. Exclusion criteria included age <1 month^14^, missing birthdate, hospital readmission, or admission during the run-in or wash-out period.

For FLUID-ICU, it is necessary to define a cohort of critically ill patients within the FLUID trial. However, the identification of a cohort of critically ill patients at true baseline (i.e., before the receipt of study fluid) in the FLUID trial is not possible since these patients would have first presented in a different geographical hospital location (ex: ED, hospital ward, operating room, post operative assessment unit) and would have very likely received an amount of NS or RL.Therefore, we used two alternative definitions for identifying the critically ill population: (1) any patient admitted to the ICU within 1 day after hospital admission (cohort 1) and (2) any patient admitted to the ICU within 2 days after hospital admission (cohort 2). Although it is recognized that admission within 1 day of hospital admission is not a conventional baseline variable, we reasoned that the acute severity of illness related to the original insult and leading to the ICU admission would likely occur within a short window after hospital admission. In a sensitivity analysis, we broadened this criterion to include patients who were admitted to the ICU within 2 days after hospital admission.

### Randomization and Intervention

For the parent FLUID trial, clusters were randomized sequentially using unrestricted randomization. The allocation sequence was computer-generated by an independent statistician at the central co-ordinating center using a permuted block design with length 2. The allocation sequence was maintained on a password-protected computer and was revealed to site investigators approximately 1 month prior to their site initiation.

During each study period, hospital staff ensured that ward stocks were at least 80% of the allocated crystalloid study fluid at all times (with non-allocated fluid <20%). For sites with electronic physician order entry, there was an automatic substitution order for the allocated study fluid each time the treating clinician ordered either RL or NS. Bedside nurses implemented an automatic substitution order in sites with manual order entry. Physicians could opt participants out of the trial if they had clinical concerns about exposure to the specific study fluid; such participants were nevertheless included in all analyses. To monitor adherence, the inventory of RL and NS was tracked daily in all hospitals.

### Data Sources and Management

All clinical data in FLUID were obtained using population-based health administrative databases at ICES as described in the main trial report.^13^ The data sets were linked using unique encoded identifiers and analyzed at ICES. Data definitions for each baseline variable and all outcome measures are explained in the data dictionary in the main trial report.

### Sample Size

The sample size for the FLUID-ICU cohort 1 is 4372 participants as per the main trial report;^13^ the sample size for the FLUID-ICU cohort 2 is 4543 participants.

### Analyses

Baseline data will be summarized as means and standard deviations (SD) for continuous variables or counts and percentages (%) for categorical variables.

All analyses will be conducted at the individual patient-level according to the intention-to-treat principle. The primary composite outcome will be analyzed using random effects logistic regression. Treatment effects comparing RL vs. NS will be expressed as odds ratios (OR) with 95% confidence intervals. The primary analytical model will include terms for treatment and period expressed as a categorical variable, and will adjust for the following *a priori* patient risk factors to improve power and efficiency: age and Elixhauser comorbidity index as continuous variables, and sex and type of admission (medical and mental health, surgical and pregnancy and childbirth), as categorical variables. The analysis will account for within-period and between-period intracluster correlation using random cluster and cluster by period interaction terms. The between-within degree of freedom method will be used to account for small sample bias^15^.

### Missing Data

Based on the main FLUID trial, we anticipate no missing outcome data and very few missing covariates. If necessary, as in the main FLUID trial, mean imputation within hospitals will be used to account for any missing baseline covariates.

### Sensitivity Analyses

As all clusters initiated the trial sequentially rather than concurrently, sensitivity analyses will be conducted, treating period as a continuous variable modelled either as a linear term or using a restricted cubic spline.

### Subgroups

The primary composite outcome will be examined for the main cohort and the sensitivity analysis cohort according to the following pre-specified subgroups of participants who are generally more likely to receive greater fluid exposure, have a higher risk of death, or adverse outcomes as described in the main trial report. These include: age (1 month to 65, > 65 to 80 and > 80 years of age); sex (male, female); co-morbid illnesses (Elixhauser Quartiles)^43^; type of hospital admission (medical and mental health, surgical and pregnancy and childbirth); sepsis (infection + organ dysfunction versus no), traumatic brain injury (yes versus no); and trauma with an injury severity score of >=12 (yes versus no). These analyses will be conducted by including the subgroup indicator as well as its pairwise interaction with period and treatment into the analytical model, with covariates as specified in the primary analytical model. Treatment effect estimates within each subgroup will be obtained from the model using least square mean differences and 95% confidence intervals and reported using forest plots with interaction p-values. Interpretation of subgroup findings will be considered exploratory, recognizing the potential for spurious findings due to multiple testing.^15^

## Data Availability

This is a secondary analysis of critically ill patients from the FLUID trial (L. McIntyre et al, N Engl J Med 2025;393:660-670).
The dataset from this study is held securely in coded form at ICES. While legal data sharing agreements between ICES and data providers (e.g., healthcare organizations and government) prohibit ICES from making the dataset publicly available, access may be granted to those who meet pre-specified criteria for confidential access, available at www.ices.on.ca/DAS.

## Ethics and Dissemination

The research ethics board approved a waiver of patient informed consent for FLUID as the intervention was cluster-level and posed minimal risk to participants^16^. The protocol and statistical analysis plan for FLUID were published^17^. FLUID was registered on clinical.trials.gov (NCT 04512950). No additional ethics review was required for this secondary analysis of anonymized data from the FLUID trial.

## Results

The results of FLUID-ICU will describe the cohorts of critically ill patients, and provide relevant current evidence about the effect of RL versus NS on clinically and patient important outcome measures, which will add to the existing literature.

## Discussion

Strengths of the FLUID-ICU study include a diverse and representative patient population of critically ill participants with heterogeneous diagnoses from 7 hospitals which enhances the generalizability of the study findings. Participants in FLUID-ICU received the allocated study fluid from hospital entry to hospital discharge, which reduces contamination with the alternate study fluid. This study will be guided by a pre-specified analysis plan and generate contemporary data related to crystalloid fluid evidence in the critically ill.

Limitations of the FLUID-ICU study include no individual participant fluid adherence data due to the infeasibility of this approach in this cluster cross-over design used for FLUID. However, adherence in the RL arm as measured by each hospital’s inventory system was somewhat lower than in the NS arm (78.2% and 93.6%). Health administrative data as compared to data collected in trials by research co-ordinators may be less accurate and prone to misclassification bias. However, misclassification in FLUID-ICU, as in FLUID would be expected to be non-differential. Finally, the complex implementation of this hospital-wide intervention necessitated sequential trial commencement in different clusters, rending the trial vulnerable to potential time-varying confounding due to seasonal effects, albeit not observed in FLUID.

## Knowledge Translation

The results of FLUID-ICU will be shared with the Canadian Critical Care Trials Group and at other national and international critical care meetings. We will present and publish our study findings in the form of abstracts, posters and peer-reviewed manuscripts. We will work with international critical care colleagues to update evidence syntheses and practice guidelines for critical care practitioners for consideration in the care of critically ill patients globally.

## Conclusions

FLUID-ICU will provide additional evidence and insights on the effects of RL compared to NS in the critically ill population. This study will also serve to generate hypotheses for future trials testing fluid strategies in the vulnerable ICU population.

